# B cell numbers predict humoral and cellular response upon SARS-CoV-2 vaccination among patients treated with rituximab

**DOI:** 10.1101/2021.07.19.21260803

**Authors:** Ana-Luisa Stefanski, Hector Rincon-Arevalo, Eva Schrezenmeier, Kirsten Karberg, Franziska Szelinski, Jacob Ritter, Bernd Jahrsdörfer, Hubert Schrezenmeier, Carolin Ludwig, Arne Sattler, Katja Kotsch, Yidan Chen, Anne Claußnitzer, Hildrun Haibel, Fabian Proft, Gabriella Maria Guerra, Pawel Durek, Frederik Heinrich, Marta Ferreira Gomes, Gerd R. Burmester, Andreas Radbruch, Mir-Farzin Mashreghi, Andreia C. Lino, Thomas Dörner

## Abstract

**Objectives:** Patients with autoimmune inflammatory rheumatic diseases receiving rituximab (RTX) therapy show substantially impaired anti-SARS-CoV-2 vaccine humoral but partly inducible cellular immune responses. However, the complex relationship between antigen-specific B and T cells and the level of B cell repopulation necessary to achieve anti-vaccine responses remain largely unknown.

**Methods:** Antibody responses to SARS-CoV-2 vaccines and induction of antigen-specific B and CD4/CD8 T cell subsets were studied in 19 rheumatoid arthritis (RA) and ANCA-associated vasculitis (AAV) patients receiving RTX, 12 RA patients on other therapies and 30 healthy controls after SARS-CoV-2 vaccination with either mRNA or vector based vaccines.

**Results:** A minimum of 10 B cells/µL in the peripheral circulation was necessary in RTX patients to mount seroconversion to anti-S1 IgG upon SARS-CoV-2 vaccination. RTX patients lacking IgG seroconversion showed reduced antigen-specific B cells, lower frequency of TfH-like cells as well as less activated CD4 and CD8 T cells compared to IgG seroconverted RTX patients. Functionally relevant B cell depletion resulted in impaired IFNγ secretion by spike-specific CD4 T cells. In contrast, antigen-specific CD8 T cells were reduced in patients independently of IgG formation.

**Conclusions:** Patients receiving rituximab with B cell numbers above 10 B cells/µl were able to mount humoral and more robust cellular responses after SARS-CoV-2 vaccination that may permit optimization of vaccination in these patients. Mechanistically, the data emphasize the crucial role of co-stimulatory B cell functions for the proper induction of CD4 responses propagating vaccine-specific B and plasma cell differentiation.

## Introduction

Infectious diseases and associated complications comprise an important cause of morbidity and mortality in patients with autoimmune inflammatory rheumatic diseases (AIIRDs) ^1^. Increased susceptibility to infectious diseases in these patients is most likely due to an immunosuppressive effect of the disease itself and/or related to immunosuppressive treatment ^2^. COVID-19, caused by the severe acute respiratory syndrome coronavirus-2 (SARS-CoV-2) requires particular considerations in AIIRD patients by rheumatologists. Rituximab (RTX), an anti-CD20 monoclonal antibody leading to B cell depletion and used in AIIRDs like rheumatoid arthritis (RA) and ANCA-associated vasculitis (AAV), has been found as risk factor for poor COVID-19 associated outcomes. ^3 4^. Since a suitable treatment for COVID-19 has not been developed yet, vaccination is of crucial importance to protect these vulnerable patients. Meanwhile, various phase III clinical trials have demonstrated the efficacy and safety of mRNA-based vaccines (BNT162b2 ^5 6^, mRNA-1273 ^7^) and viral vector-based vaccines (ChAdOx1 ^8^, Ad26.COV2.S ^9^) to prevent severe COVID-19 disease or death.

In AIIRD patients, vaccination is generally regarded safe and efficacious ^10^. However, in particular under B cell depleting therapy with RTX, hampered humoral and cellular responses following influenza, pneumococcal and hepatitis B vaccination have been reported ^11-16^. Data available about SARS-CoV-2 vaccine response in rituximab treated AIIRD patients reveal substantially impaired humoral ^17-19^ but partly inducible cellular immune responses ^20^. However, little is known about the complex mechanisms between T, B and plasma cells, as well as the level of B cell repopulation necessary for proper vaccine response among RTX patients.

In this study, we investigated the characteristics of humoral and cellular antigen-specific CD4/CD8 and B cell immune response upon SARS-CoV-2 vaccines in patients treated with RTX compared with HC and RA patients on other therapies.

## Results

### Cohorts and patient characteristics

This study recruited 19 patients receiving rituximab (16 RA and 3 AAV patients, RTX group), 30 healthy controls (HC group) and 12 RA patients on other therapies as additional control group (RA group). Most study participants were vaccinated twice with the mRNA vaccine BNT162b2, one RTX patient received 2x mRNA-1273. There were 3 HC, one RA and one RTX vaccinated twice with the viral vector vaccine ChAdOx1 (indicated in green throughout the figures). According to national recommendations ^21^, 3 RTX patients and 3 HC received 1x ChAdOx1 followed by a heterologous boost with 1x BNT162b2 (indicated in blue throughout the figures). Regarding demographics, HC were younger than RA patients, while age-matched to RTX patients. As known for patients with rheumatic diseases, the majority of RA and RTX patients were female. Disease activity at the time of first vaccination was comparable between the RA control group and RA patients in the RTX group. At the time of vaccination, median time since the last RTX treatment was 9 months. RTX patients had received B cell depleting therapy on average for 3 years, presenting with a range of circulating B cell number between 0 - 484/µl blood. Demographics and co-medication of all patients are summarized in Table 1.

**Table 1:** Patient characteristics.

|  | HC<br>N=30 | RA<br>N=12 | RTX<br>N=19 (16 RA, 3 AAV) |
| --- | --- | --- | --- |
| **Age** |  |  |  |
| Median [IQR] | 57<br>[46.25 – 79.5] | 68<br>[63 – 79.5] | 58<br>[56.5 - 65] |
| Under 50 | 9 | 1 | 3 |
| Between 50-69 | 12 | 5 | 13 |
| > 70 | 9 | 6 | 3 |
| **Gender** |  |  |  |
| Female | 15 | 9 | 14 |
| Male | 15 | 3 | 5 |
| **Vaccines (n)** |  |  |  |
| 2x BNT162b2 | 24 | 11 | 14 |
| 2x mRNA-1273 | 0 | 0 | 1 |
| 2x ChAdOx1 | 3 | 1 | 1 |
| 1x ChAdOx1, 1x BNT162b2 | 3 | 0 | 3 |
| **Immunosuppression (n)** |  |  |  |
| MTX |  | 8 | 4 |
| Leflunomid |  | 1 | 0 |
| Sulfasalazin |  | 0 | 1 |
| AZA |  | 0 | 1 |
| JAKI |  | 4 | 2 |
| TNFI |  | 1 | 0 |
| Abatacept |  | 2 | 1 |
| Prednisolone |  | 3 (max 5mg/d) | 8 (max 7.5mg/d) |
| **DAS 28** |  |  |  |
| Median [IQR] |  | 2.58 [1.70 – 3.99] | 2.65 [1.96 – 3.41] |
| **Months since last RTX** |  |  |  |
| Median [IQR] |  |  | 9 [6 – 13.5] |
| **Years on RTX** |  |  |  |
| Median [IQR] |  |  | 3 [2 - 6] |

### Impaired humoral response upon SARS-CoV-2 vaccination in RA and RTX patients

Antibody responses to SARS-CoV-2 vaccines were assessed in all individuals, 6±3 days after boost. All HC became positive for anti-S1 IgG and IgA and showed more than 90% SARS-CoV-2 neutralisation. Noteworthy, IgA and IgG anti-vaccine titres were markedly diminished 6±3 days after boost in the RA control group and especially in RTX patients compared to HC (Fig. 1A). Anti-S1 IgG were detected in 8/12 (66.7%) of the RA group and 8/19 (42.1%) of the RTX group. Simultaneously, 5/12 (41.7%) and 9/19 (47.4%) of the RA and RTX group, respectively developed anti-S1 IgA antibodies. Virus neutralizing antibodies were found in 8/12 (66.7%) among RA control patients and 9/19 (47.4%) RTX group (Fig. 1A).

**Figure 1:**
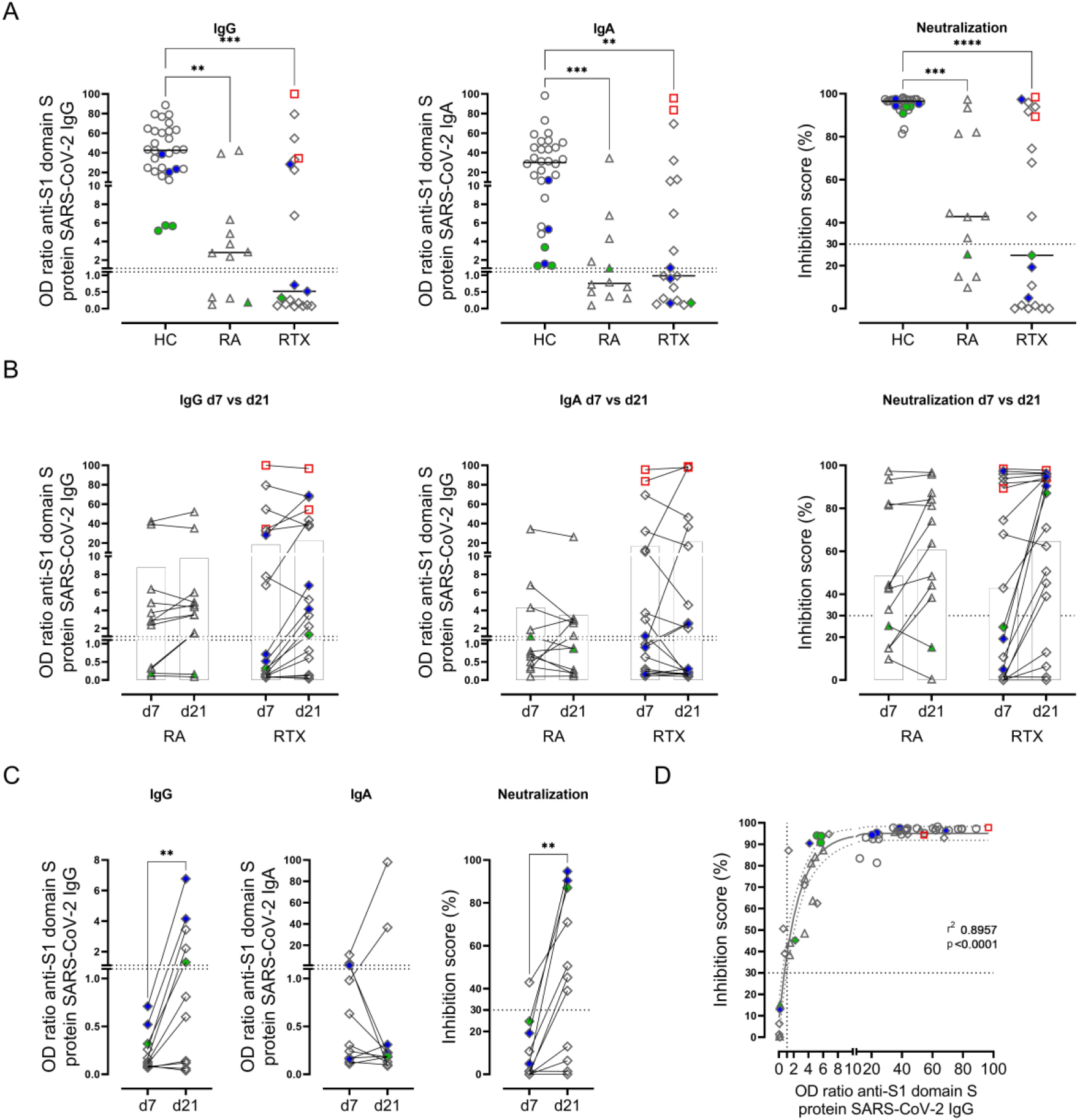
Humoral immune response is reduced and delayed in RA and RTX patients. (A) Humoral immune response against SARS-CoV-2 was assessed by ELISA for spike protein S1 IgG, spike protein S1 IgA and virus neutralization by a blocking ELISA in HC (n=30), RA (n=12) and RTX (n=19) 6±3 days after 2^nd^ SARS-CoV-2 vaccination. Threshold of upper limit of normal is indicated by dotted lines. (B) Follow-up sera were collected for 12 RA and 19 RTX patients, respectively, 3-4 weeks after 2^nd^ vaccination and investigated for S1 IgG, IgA and virus neutralization. (C) Delayed IgG response in 5/11 of initially (day 7 after boost) non-seroconverted RTX patients at day 21. (D) Significant correlation between IgG titers and neutralizing antibodies among RTX patients. (A) Kruskal-Wallis with Dunn’
ss post-test. (B) Two way ANOVA with šidák post-test. Interaction effect was not significant. (C) Mann Whitney test. (D) Spearman correlation. Sigmoidal model with 95% confidence bands. *p<0.05, **p<0.01, ***p<0.001, ****p<0.0001. Color code: previously infected individuals are indicated as red quadrats; 2x vaccinated with ChAdOx1 indicated in green; 2x heterologous vaccinated 1x ChAdOx1 followed by 1x BNT162b2, indicated in blue.

Two RTX patients with unknown prior infection (identified as anti-nucleocapsid protein positive Fig. S1, red quadrates), developed high titers of anti-S1 IgG, IgA and neutralizing antibodies comparable with HC.

### Delayed serologic response occurred in RTX patients

As previously reported ^22^, AIIRD patients may show a delayed humoral immune response after vaccination. To address this question, we collected additional samples from RA and RTX patients 3-4 weeks after boost (Fig. 1B). Two RA and five RTX patients developed positive IgG antibodies, IgA was found in two RA and one RTX patients 3-4 weeks after boost. Neutralizing antibodies were detected in two RA and six RTX at this later time point. Among the RTX patients, who did not show seroconversion at day 7 after boost, there was a significant increase in IgG and neutralizing antibody formation at the later time point (Fig. 1C). Noteworthy, IgG titers correlated with neutralizing antibodies (Fig. 1D). Thus, and considering the delayed vaccine response, 10/12 (83.3%) of RA patients and 13/19 (68.4%) of the RTX patients showed IgG seroconversion with neutralizing antibodies after SARS-CoV-2 vaccination, even though in lower titers compared with HC. Interestingly, the data suggested that patients under RTX exhibited a potential dichotomous response: 13/19 RTX patients seroconverted to IgG (RTX IgG+), while 6/19 did not (RTX IgG-). To identify potential factors resulting in IgG seroconversion among RTX patients, further study addressed potential differences between the two groups.

### A minimum of 10 B cells/µl appeared to be required for specific IgG induction

We analyzed the B cell compartment among the different groups (gating strategy shown in Fig. 2A). RTX patients presented with significantly lower relative and absolute B cell numbers compared to HC and RA control group (Fig. 2B). Notably, a significant difference in the frequency and absolute B cell number was also found between RTX IgG+ and non-seroconverted RTX patients (Fig 2C). In our RTX cohort, 10 B cells/µl in peripheral circulation (or 0.4% of lymphocytes accordingly) were identified as the minimum to mount seroconversion to anti-S1 IgG among RTX treated patients (Fig 2C).

**Figure 2:**
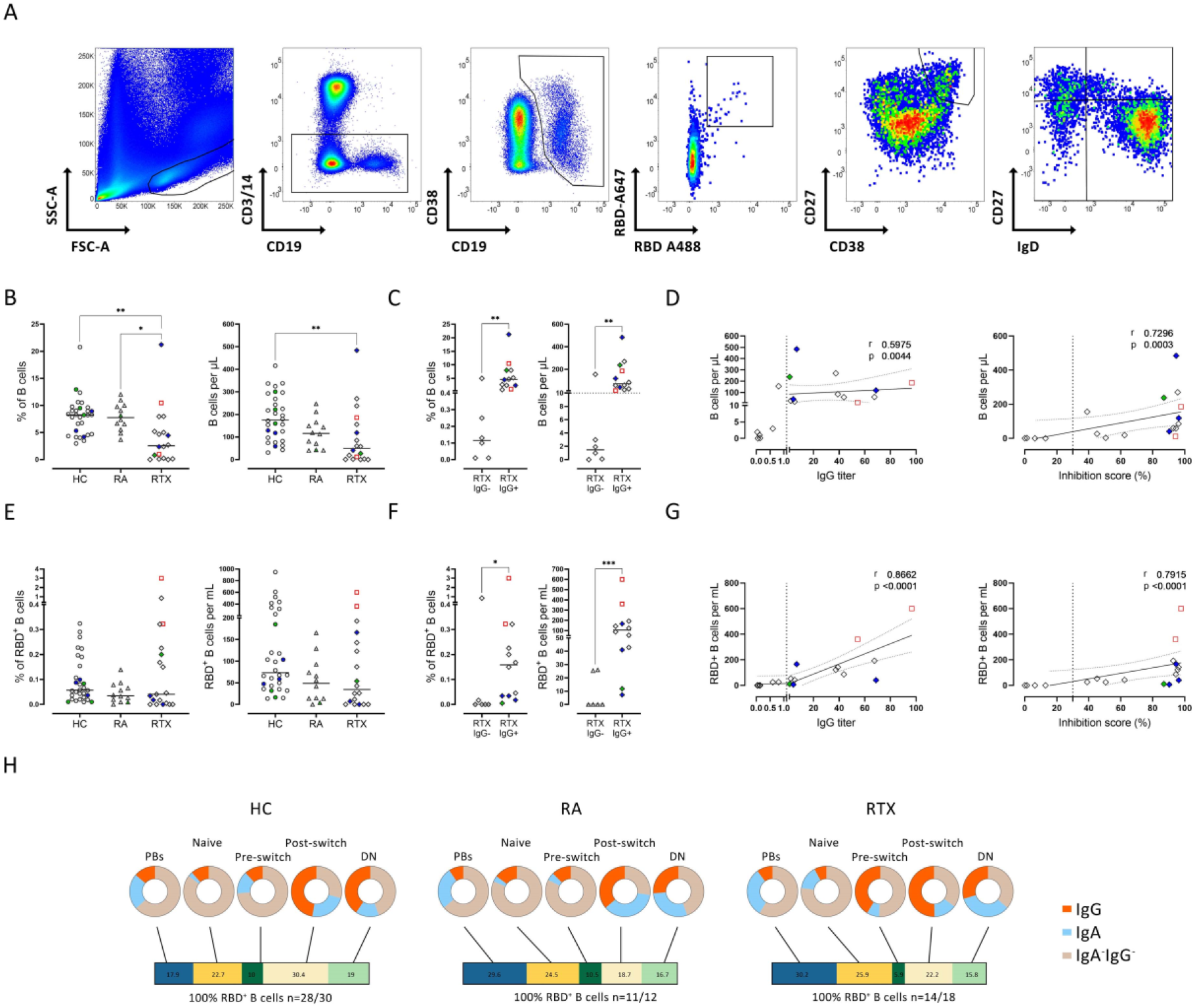
Total B cells and antigen specific B cells are significantly reduced in RTX patients and correlates with IgG formation and neutralizing capacity. (A) Representative pseudocolor plots of lymphocytes, CD3-CD14-cells, CD19+ B cells, RBD+ B cells, plasmablasts, and non-plasmablast B cell subsets based on IgD/CD27 classification. Frequency and corresponding absolute number (measured by BD Trucount) of CD19+ B cells in (B) HC (n=30), RA (n=12) and RTX (n=18) and in (C) RTX IgG+ (n=12) compared to RTX IgG- (n=6) 6±3 days after 2^nd^ vaccination. (D) Direct correlation between CD19+ B cells and IgG formation as well as neutralizing capacity in RTX patients. Frequencies and absolute numbers of RBD+ cells among total CD19+ B cells measured 6±3 days after 2^nd^ vaccination in (E) HC, RA and RTX and (F) in RTX IgG+ and IgG-. (G) Direct correlation between RBD+ B cells and IgG formation as well as neutralizing capacity in RTX patients. (H) Frequencies of plasmablasts, naïve, pre-switch, post-switched and double negative RBD+ B cells (bar) and immunoglobulin isotype distribution among subsets (cakes) in HC, RA and RTX patients, 6±3 days after 2^nd^ vaccination. Only donors with detectable RBD+ B cells were included: HC (n=28/30), RA (n=11/12) and RTX (n=14/18). (B-C, E-F) Kruskal-Wallis with Dunn’
ss post-test. (D, G) Spearman’s correlation. Linear model with 95% confidence bands. *p<0.05, **p<0.01, ***p<0.001, ****p<0.0001. Spearman’s correlation. Color code: previously infected individuals are indicated as red quadrats; 2x vaccinated with ChAdOx1 indicated in green; 2x heterologous vaccinated 1x ChAdOx1 followed by 1x BNT162b2, indicated in blue.

In the RTX group, B cell numbers correlated with humoral anti-vaccine responses since the absolute number of B cells (Fig. 2D) and B cell frequency (Fig. S2A) correlated with anti-S1 IgG titers and even more pronounced with neutralizing antibodies. This clearly suggests that humoral protection elicited by vaccination is dependent on the critical availability of B cells in RTX treated patients. In the RA and HC groups we did not find a significant correlation between B cell numbers and serologic response (data not shown), suggesting that the correlation between B cell numbers and IgG response is restricted to patients with B cell counts below the lower limits of normal.

### Vaccine non-responders among RTX patients show significant lower antigen-specific B cells

Next, we studied SARS-CoV-2 specific B cell responses in RTX treated patients, using flow cytometry to quantify RBD-specific B cells in peripheral blood ^23^ (gating strategy shown in Fig 2A). While no significant difference was seen between HC, RA and the RTX group (Fig. 2E), RTX patients lacking seroconversion upon boost (RTX IgG-) showed significantly reduced frequencies and absolute numbers of RBD+ specific B cells compared with RTX IgG+ patients (Fig. 2F). The number (Fig. 2G) and frequency (Fig. S2A) of RBD+ B cells in RTX patients correlated with the induction of IgG and neutralizing antibodies. Subsequent analyses addressed the distribution of RBD-specific B cells among B cell subsets. As previously shown for HC ^23^, RA control and RTX patients were able to generate IgG+ plasmablasts upon vaccination. We found no significant difference between the groups regarding RBD+ B cell subset distribution or immunoglobulin isotypes (Fig. 2H and S2B, C).

### TfH-like and activated CD4 and CD8 T cells are reduced in IgG non-responders under RTX treatment

Simultaneously, we wondered how dynamics of CD4/8 T cell subsets interrelate with induction of vaccine-specific B cells and IgG. Contrary to B cells, there was no difference regarding the frequency, absolute numbers or memory formation in CD4 (Fig. S3A, C) and CD8 T cells (Fig. S3B, D) between HC, RA and RTX patients. Subsequent analysis addressed the differences between vaccine-responders and non-responders among RTX patients (representative gates shown in Fig. 3A). Interestingly, patients who lacked anti-vaccine IgG antibodies showed lower frequencies of circulating TfH-like CD4 T cells, defined as CD4+CXCR5+PD1+, as well as of activated CD4/8 T cells co-expressing CD38+HLA-DR+ (Fig. 3B). Activated CD4 T cells correlated with absolute B cell numbers (Fig. 3C). This data suggests an impaired bidirectional T-B cell interaction in patients with gradual B cell depletion that results in insufficient vaccination-induced humoral immunity.

**Figure 3.**
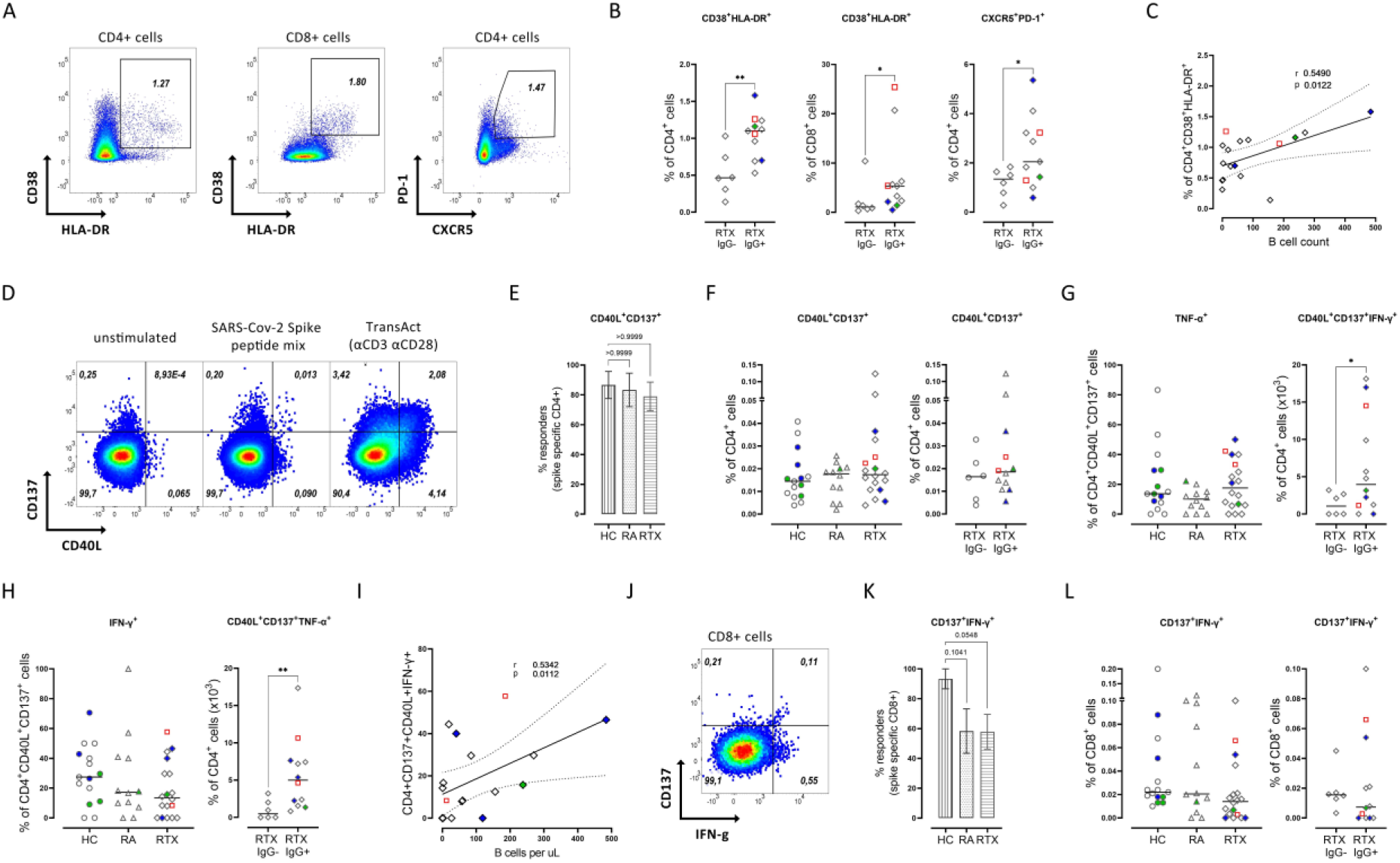
Activated CD4 T cells and IFNγ+ antigen-specific CD4+ T cells correlate with absolute B cell counts. (A) Representative plots for activated, CD38+HLA-DR+ CD4 and CD8 T cells and CXCR5+PD-1+ TfH-like CD4 T cells. (B) Activated CD4, CD8 T cells as well as circulating TfH-like CD4 T cells are significantly lower among vaccine non-responders (n=6) compared to responders (n=11, due to partial missing data in 1 RTX patient) of the RTX cohort. (C) Activated CD4 T cells correlates with B cell count. (D) Representative plots for antigen-specific CD4 T cells (CD137+CD40L+) after peptide stimulation compared with no stimulation and TransAct stimulation. (E) Responder rate and (F) frequency of antigen-specific CD4 T cell induction with B.1.1.7 SARS-CoV-2 Spike peptide mix stimulation; (G) TNFα+ and (H) IFNγ+ production of spike-specific CD4+ T cells after induction with B.1.1.7 SARS-CoV-2 Spike peptide mix stimulation in HC (n=15), RA (n=12), and RTX (n=18), and in RTX IgG+ (n=12) and IgG- (n=6). (I) Correlation between IFNγ+ antigen-specific CD4 T cells and B cell count. (J) Representative plot for antigen-specific CD8 T cells (CD137+IFNγ+). (K) Responder rate and (L) frequency of antigen-specific CD8+ T cells after induction with B.1.1.7 SARS-CoV-2 Spike peptide mix stimulation. (B, F-H, L) Mann Whitney test. (E-H, K-L) Kruskal-Wallis with Dunn’
ss post-test. (C, I) Spearman correlation. Linear model with 95% confidence bands. *p<0.05, **p<0.01, ***p<0.001, ****p<0.0001. Color code: previously infected individuals are indicated as red quadrats; 2x vaccinated with ChAdOx1 indicated in green; 2x heterologous vaccinated 1x ChAdOx1 followed by 1x BNT162b2, indicated in blue.

### Impaired cytokine secretion of antigen-specific CD4 T cells is characteristic of RTX IgG-patients and correlates with absolute B cell number

The overall occurrence of spike-specific CD4 T cells (representative gates shown in 3D) compared with unstimulated controls was found similar in all groups: 86.7% (13/15) in HC, 83% (10/12) in RA and 73.7% (14/19) in RTX patients (Fig 3E). This was also consistent with a comparable magnitude of the response between the groups (Fig. 3F) as well as similar memory subset distribution (Fig. S4C). A more detailed study of the RTX group showed that the majority of RTX IgG+ patients (10/13, 76.9%) versus 50% of RTX IgG- (3/6) patients showed an appropriate antigen-specific CD4 T cell increase upon stimulation. With regard to functional analyses of cytokine secretion by spike-specific CD4 T cells, non-seroconverted RTX patients showed a significantly reduced TNFα (Fig 3G) and IFNγ (Fig. 3H) production compared to RTX IgG+ responders (representative gates shown in Fig. S4A).

Since most patients in the non-seroconverted RTX group had very low circulating B cell counts, we wondered if there is a relation between reduced B cells and impaired cytokine production by antigen-specific CD4+ T cells. Indeed, IFNγ but not TNFα production showed a correlation with absolute B cell number, suggesting the importance of B cell co-stimulatory functions for the proper and interactive induction of CD4 responses.

### Lower antigen-specific CD8 responses in RA and RTX patients

Compared with unstimulated controls, 93.3% of HC (14/15) but only 58.3% of RA control (7/12) and 57.9% of RTX patients (11/19) showed an increase of spike-specific CD8 T cells co-expressing CD137 and IFNγ (as shown in Fig. 3J and Fig. S4B) upon stimulation (Fig. 3K). To assess the degranulation function of CD8 T cells, we analyzed the co-expression of CD107a and IFNγ. The responder rate for CD8 T cells co-expressing CD107a and IFNγ after stimulation was overall low with 60% in HC (9/15), 41.6% (5/12) in the RA control and 42.1% (8/19) in the RTX group (data not shown). Regarding the amplitude of CD8 responses, spike-specific CD8 T cells, co-expressing CD137 and IFNγ (Fig. 3L) and CD107a and IFNγ (Fig. S4C), as well as their memory subset distribution were comparable between all groups (Fig. S4D-E).

### Antigen specific and activated T cell subsets correlate with RBD+ plasmablasts

To identify further predictive factors for IgG seroconversion in RTX patients, we performed a correlation matrix (Fig. 4) including antigen-specific T and B cell subsets as well as demographic data. IgG titers and neutralizing antibodies correlated with RBD+ plasmablasts and memory compartments, as we previously showed ^23^. Furthermore, neutralizing antibodies correlated with the frequency of activated CD38+HLA-DR+ CD4/8 T cells as well as with IFNγ producing antigen-specific CD4 T cells. Activated CD38+HLA-DR+ CD4/8 T cells correlated also with RBD+ plasmablasts, while circulating TfH-like CD4 T cells with total RBD+ B cells. Notably, there was a correlation between TNFα and IFNγ producing antigen-specific CD4 T cells with RBD+ plasmablasts and switch-memory B cells. There was a correlation between IgG titer and interval of time since the last RTX infusion, but no significant correlation with age or with DAS28.

**Figure 4.**
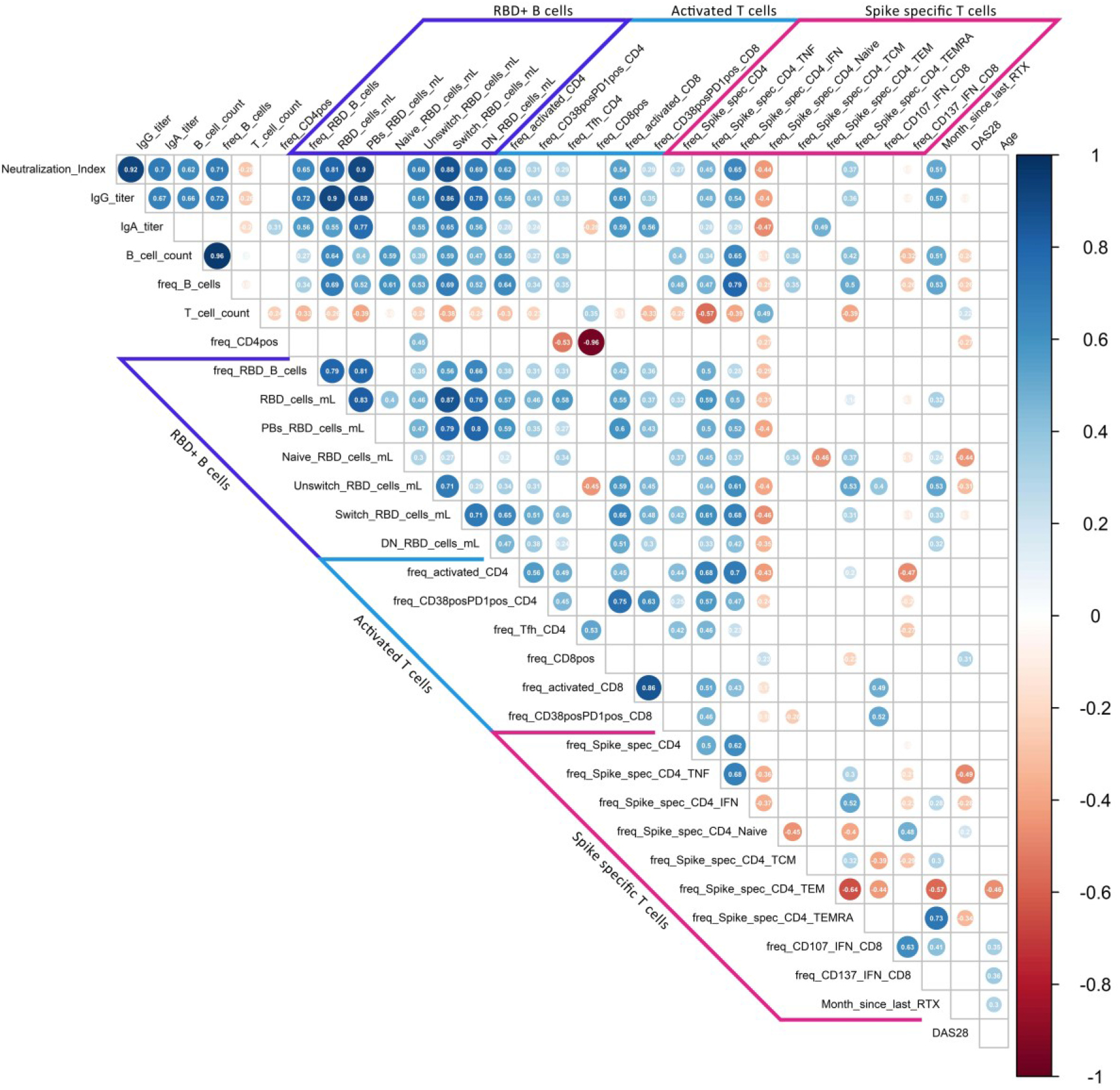
Correlation of humoral and cellular vaccine responses in RTX treated patients. Spearman’
ss correlation matrix showing relation between humoral responses, RBD+ B cell subsets, activated CD4 and CD8 T cells, antigen-specific CD4 and CD8 T cell response, and demographics. Corresponding correlations are represented by red (negative) or blue (positive) circles; size and intensity of color refer to the strength of correlation (RTX n=17 due to partial missing data in 2 RTX patients). Only correlations with p ≤ 0.05 are indicated. Indicated values in circles indicates r value of correlation.

Interestingly, induced antigen-specific CD8 responses upon stimulation did not correlate with humoral immunity, neither with B nor CD4 T cell subsets, suggesting an independent, more direct antigen-driven cellular immunity compared with CD4/CD19-interaction required for IgG formation.

### Circulating follicular B cells are reduced in RTX IgG-patients

To further investigate the specific differences during SARS-CoV-2 vaccination in RTX treated patients, we sorted CD27++CD38++ plasmablasts, CD27+ memory B cells and HLA-DR+CD38+ activated T cells as indicators of the ongoing adaptive immune response after vaccination. The cytometrically enriched cells were subsequently analyzed using DropSeq single cell RNA sequencing technique (^23^). Unsupervised analysis using Uniform Manifold Approximation and Projection for Dimension Reduction (UMAP) could identify 15 distinct cluster: 4 B cell, 3 plasmablast and 8 clusters of activated T cells (Fig. 5A-C). Here, clusters 3 and 5 were of particular interest: cluster 3 is enriched with circulating follicular-like B cells expressing *CXCR5* and *CCR6* and cluster 5 contains *CD40LG, PDCD1* and *ICOS* expressing TfH-like CD4 T cells. Follicular B cells and *CD40LG+PD1+* Tfh cells were substantially reduced in RTX patients and most pronounced among IgG non-responders in the RTX group (Fig. 5 D).

**Figure 5.**
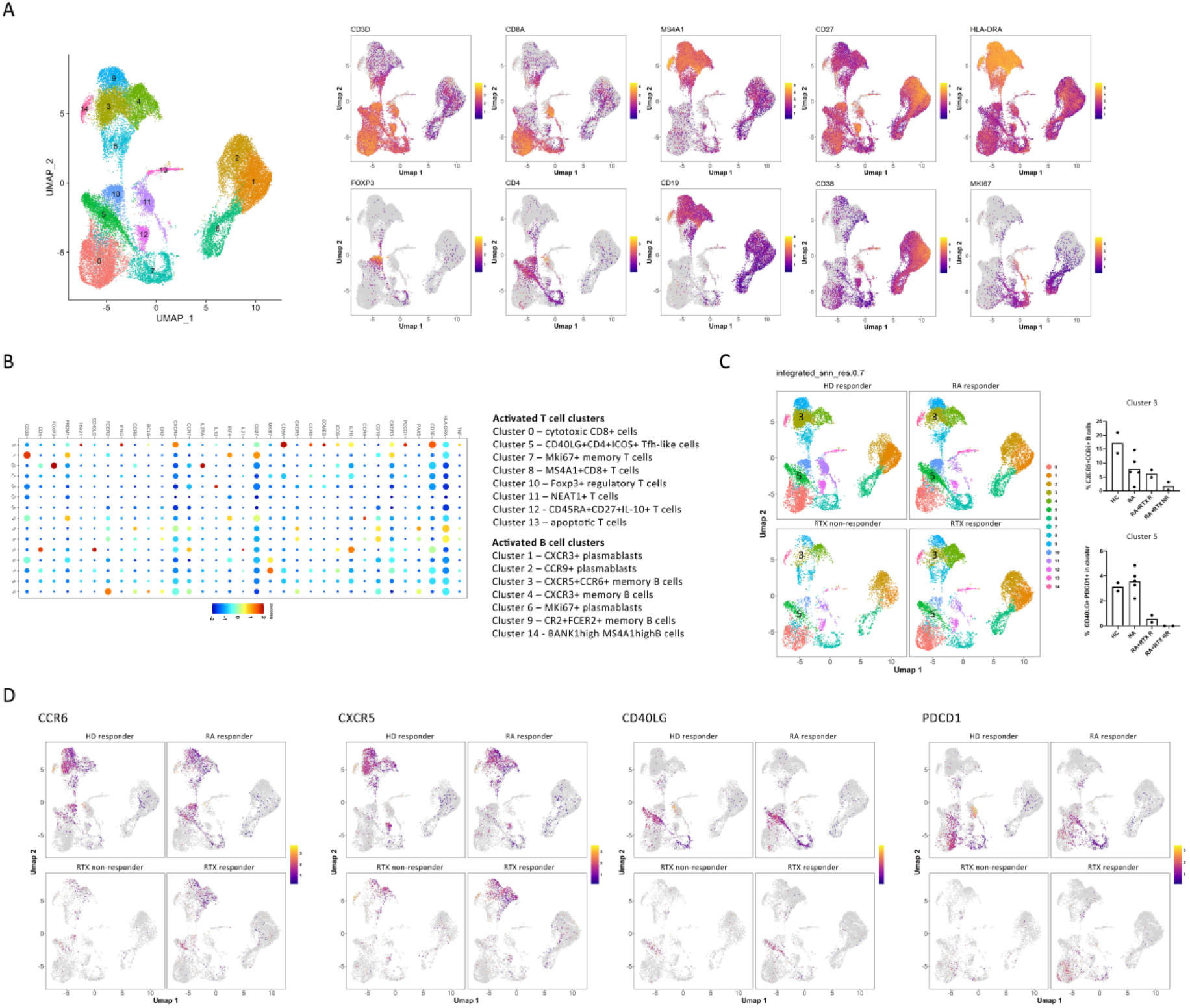
Single cell transcriptome and CITE Seq analyses reveal diminished follicular T and B cells in RTX patients. (A) UMAP clustering of peripheral blood CD27++CD38++ plasmablasts, CD27+ memory B cells and T cells from 2HC, 4 RA and 5 RTX patients (3 responders and 2 non-responders) were isolated and sorted by FACS for single cell sequencing. (B) Relative expression levels of selected signature genes in the 13 identified clusters (total cell number sequenced 38038). (C) UMAP clustering from 2 HC, 4 RA, 3 RTX IgG+ and 2 RTX IgG-patients and cluster frequency comparison for cluster 3 and 5. (D) UMAP representation of the expression levels of *CCR6, CXCR5, CD40LG* and *PDCD1* in HC, RA, RTX IgG+, and RTX IgG-patients.

## Discussion

SARS-CoV-2 vaccines have been approved based on their protection against COVID-19 in clinical trials ^5-8^. However, certain patient groups receiving immunosuppressive therapies appear to develop insufficient humoral and cellular responses ^18 23-25^, but limited data about the underlying limitations is available. Protection through immunization is achieved by an orchestrated immune response between different cellular subsets of innate (APCs) and adaptive immunity, such as B and T cells. Anti-B cell therapies like anti-CD20 antibodies (rituximab, obinutuzumab and ocrelizumab) and BTK inhibitors are associated with poor humoral SARS-CoV-2 vaccination responses, in patients with AIIRD ^17-20^, multiple sclerosis ^26^ and CLL ^27^. Since B cell depletion enhances the risks for poor Covid-19 outcomes ^3^, but also can reduce anti-SARS-CoV-2 vaccine responses, it is of utmost importance to delineate the level of B cell repopulation necessary to achieve anti-vaccine responses and get insights into the complex relationship between antigen-specific B and T cells.

Therefore, our study aimed to investigate humoral and cellular responses in RTX treated patients versus controls. Consistent with previous data ^17-19 22^, serologic IgG conversion with formation of neutralizing antibodies was significantly lower and delayed in both, RA and even more pronounced in the RTX cohort compared to HC. This finding was closely linked to the availability of peripheral B cells, activated CD4/8 T cells as well as circulating TFH-like cells. Ongoing antigen exposure through mRNA vaccines seems to permit prolonged GC maturation ^28^, which might be an explanation for the further increase in antibody titers during an additional period in some patients.

Besides IgG non-responders among RTX patients, two patients in the RA group with normal B cell numbers did not develop anti-S1 IgG antibodies. After completing the analysis, the underlying cause may be most probably related to impaired T cell responses: in one patient due to inhibition of co-stimulation by abatacept, consistent with a prior report ^18^. The other patient treated with a JAK inhibitor lacked cytokine production by antigen-specific T cells after ChAdOx1 booster. Even though significantly lower IgG responses were reported upon ChAdOx1 booster in healthy vaccinees ^29^, it remains to be delineated whether the treatment and/or selected vaccine may account for this finding. Induction of vaccine-specific IgG in individuals upon ChAdOx1/BNT162b2 was comparable with twice BNT162b2 vaccinations. Interestingly, IgA formation was comparable across all groups, although the protective potency of IgA remains to be determined.

Of utmost importance, our RTX cohort showed a correlation between IgG seroconversion, neutralizing antibodies and absolute B cell number. Here, a minimum of 10 B cells/µl in the peripheral circulation appeared to be a threshold to signify an appropriate cellular and humoral vaccination response. Patients with B cell numbers below this threshold presented not only with lower antigen-specific B cells, but also they showed substantially diminished circulating TfH-like CD4 T cells, reduced activated CD4/8 T cells co-expressing CD38 and HLA-DR, as well as impaired IFNγ secretion of spike-specific CD4 T cells. The frequency of IFNγ secreting antigen-specific CD4 T cells also correlated with the absolute number of B cells, suggesting that these cells interact to achieve proper anti-S1 responses. Mechanistically, the current data suggest the critical role of available co-stimulatory B cell functions for the induction of proper CD4 Th response. This is consistent with observations of previously described impaired B-T cell crosstalk in rituximab treated patients ^30-32^, leading to reduced frequencies of activated T cells ^32^, downregulation of CD40L in CD4 T cells ^30 31^, and reduced antigen-specific CD8 T cells after influenza vaccination ^14^.

With regard to induction of antigen-specific CD8 T cells upon stimulation, the RTX and RA groups showed a reduced responder rate compared with HC. However, other than for antigen-specific CD4 T cells, neither B cell depletion nor IgG formation correlated with spike-specific CD8 T cells, suggesting that their induction occurred independently upon SARS-CoV-2 vaccination. It is not clear how these vaccine-specific CD8 T cells provide antiviral protection on clinical grounds.

The debate about what correlates with protection after vaccination against SARS-CoV-2 is ongoing, while it is widely accepted, that neutralizing antibodies are considered to be a reliable surrogate of protection against virus variants ^33 34^. The threshold for protective SARS-CoV-2 IgG-titer is still unknown, although non-human primate studies suggest that it is likely very effective already at low titers ^35^. Our study provides evidence that detection of RBD-specific B and spike-specific CD4 T cells may provide cellular correlates of this response, while the CD8 response occurred in an independent way. The role of these two lines of vaccine response needs to be further delineated.

Limitations of the study are the small number of RA and RTX patients and the heterogeneity of the groups (including different DMARD regimes, different vaccination strategies). However, co-medication with prednisolone or methotrexate do not appear to bias our results, since there was no difference in humoral response in our RTX cohort upon vaccination (Fig. S7A, B).

Here, we present a first study investigating humoral as well as antigen-specific T and B cellular responses in RTX treated patients after SARS-CoV-2 vaccination. Mechanistically, the data provide insights into the crucial role of available B cells equipped with proper co-stimulatory function to interactively cross-talk with CD4 T cells. These functions likely result into GC formation, plasma cell differentiation and vaccine-specific IgG production. As a clinical consequence, we propose a threshold of absolute B cells signifying expansion of vaccine responses after RTX treatment, which may support optimization of vaccination protocols among this vulnerable patient group.

## Data Availability

All data is available.

## Acknowledgements

We kindly thank all study participants for taking part and all team members of the rheumatology outpatient clinic Steglitz (Dr. Kirsten Karberg and Dr. Henning Christian Brandt), Berlin, as well as of AGZ rheumatology Charité Mitte Berlin (Dr. Anne Claußnitzer), Germany for organization of patient schedules.

## Author contributions

The concept of the study was developed by ALS, HRA, ES, ACL, and TD.

Patient’s samples were collected by KiK, ALS, JR, AC, HH, FP.

Data were obtained by ALS, HRA, FSZ, JR, YC, BJ, HS, CL, MFG, MFM. Data were analyzed by HRA, ALS, MFG, MFM, ACL, DP, FH, GMG.

The theoretical framework was developed by TD, ALS and HRA. The work was supervised by ACL, AS, KK, AR, GRB, and TD.

All authors developed, read, and approved the current manuscript.

## Key messages (5 bullets)

### What is already known about this subject?

- Patients on rituximab therapy are on higher risk for poor COVID-19 outcomes and show substantially impaired IgG responses after 1SARS-CoV-2 vaccination.

### What does this study add?

- The study found that B cell numbers correlate with formation of anti-vaccine neutralizing IgG antibodies. Patients having 10 B cells/µl of blood are able to mount humoral protection.
- Mechanistically, we show that lack of B cells leads to an impaired function of antigen-specific CD4 T cells.

### How might this impact on clinical practice or future developments?

- Our data support that rheumatologists can optimize SARS-CoV-2 vaccination among rituximab treated patients by monitoring B cells.
- Future treatment developments targeting T cells may take into account the directive role of critical B cell counts,

