## Supplemental data for "B cell numbers predict humoral and cellular response upon SARS-CoV-2 vaccination among patients treated with rituximab"

**Supplementary material**

**Material and Methods**

**Study participants**

Outpatient rheumatic patients treated with rituximab (from clinics in Steglitz, Charité Mitte, Charité Campus Benjamin Franklin), who received SARS-CoV-2 vaccination according to federal and Berlin state recommendations between February and May 2021, were asked to participate into this study. We included 16 RA patients (according 2010 Rheumatoid Arthritis Classification Criteria [1]) and 3 AAV patients (defined as [2]) on RTX treatment as well as 12 RA patients on other therapies and 30 HC as control groups. HC and RTX groups were age-matched to avoid potential confounders. All participants gave written informed consent according to the approval of the ethics committee at the Charité University Hospital Berlin (EA2/010/21, EA4/188/20). Peripheral blood samples (EDTA anti-coagulated or serum-tubes, BD Vacutainersystem, BD Diagnostics, Franklin Lakes, NJ, USA) were collected at 6 ± 3 days after vaccination with either 2x SARS-CoV-2 BNT162b2, 2x ChAdOx1 nCoV-19 or 1x ChAdOx1 nCoV-19 followed by 1x SARS-CoV-2 BNT162b2. Serologic and B cell data of HC have partially been published [3]. Samples 3-4 weeks after boost were available for 19 RTX and 12 RA patients. 1 RTX patient (with RA) had a concomitant chronic lymphocytic leukaemia, cellular data of this patient was excluded in the figures 2 and 3. Donor information is summarized in Table 1.

**Patient involvement**

Patients were not involved in the design and conduct of this research. The results are going to be communicate to the patients by their rheumatology specialist. We aim to publish a short communication in the newspaper of the German Rheuma Liga.

**Enzyme-linked immunosorbent assay and surrogate SARS-CoV-2 neutralization test**

Serum tubes were centrifuged at 3000rpm for 10 minutes to separate plasma. Serum was stored at -20C° prior to antibody analysis. The anti-SARS-CoV-2 assay (Euroimmun, Lübeck, Germany) is a classical enzyme-linked immunosorbent assay (ELISA) for the detection of IgG to the S1 domain of the SARS-COV-2 spike (S) protein, IgA to the S1 domain of the SARS-COV-2 spike protein, and IgG to the SARS-CoV-2 nucleocapsid (NCP) protein. The assay was performed according to the manufacturer´s instructions, as described [3]. The blocking ELISA (GenScript) qualitatively detects anti-SARS-CoV-2 antibodies suppressing the interaction between the receptor binding domain (RBD) of the viral spike glycoprotein (S) and the angiotensin-converting enzyme 2 (ACE2) protein on the surface of cells as described previously [3]. To identify previously SARS-CoV-2 infected individuals we measured antibodies against the nucleocapsid protein (NCP, not a vaccine component) 6±3 days after boost (Fig. S1).

### Isolation of peripheral blood mononuclear cells (PBMCs) and staining

PBMCs were prepared by density gradient centrifugation using Ficoll-Paque PLUS (GE Healthcare Bio-Sciences, Chicago, IL, USA). For antigen-specific T cell analysis PBMCs were cryopreserved at -80°. For surface staining 1-3 x $10^6$ cells were suspended in 50 µl of PBS/0.5% BSA/EDTA and 10 µl Brilliant Buffer (BD Horizon, San Jose, CA, USA). Cells were stained for 15 min on ice and washed afterwards with PBS Dulbecco containing 1% FCS (fetal calf serum, Biowest, Nuaillé, France) (810 xg, 8 min, 4°C). For intracellular staining after T cell stimulation cells were first stained for 30 min with 1:1000 BUV395 Life/Dead (Invitrogen) in PBS, followed by 5 min 2.5 µl Fc Block (Milteny Biotech) in 50 µl resuspended cells. Cells were fixed in LyseFix (Becton Dickinson), permeabilized with FACS Perm II Solution (Becton Dickinson) and intracellularly stained. Gating of flow cytometric analysis was performed as indicated in Figure 2 and 3.

### Staining of antigen-specific B cells

To identify RBD-specific B cells, recombinant purified RBD (DAGC149, Creative Diagnostics, New York, USA) was labeled with either AF647 or AF488 as reported [3]. Double positive cells were considered as antigen-specific. Antigens were labeled at the German Rheumatism Research Centre (DRFZ), Berlin with NHS esters conjugation for AF647 and AF488.

### Peptide stimulation for antigen specific T cells

2x$10^6$ PMBC per stimulation of 15 HC, 12 RA control and 19 RTX patients were thawed and washed twice in pre-warmed RPMI1640 medium (containing 0.3 mg/ml glutamine, 100 U/ml penicillin, 0.1 mg/ml streptomycin, 10% FCS and 25 U/ml DNase I (Roche International)), rested for 1 h in culture medium (RPMI1640 with glutamine, antibiotics and 10 % FCS) and stimulated with SARS-CoV2 spike (“PepMix” SARS-CoV-2 (S B.1.1.7), JPT, Berlin, Germany) [4] or T cell TransAct (Miltenyi Biotech) in the presence of APC anti-CD107a (Biolegend, clone H4A3) for 16 h. Brefeldin A (10 µg/ml, SigmaAldrich) was added after 2 h. Due to cell number limitations, T cell TransAct stimulation was not carried out for all particpitants. CD4 T cells co-expressing CD154 and CD137 were considered antigen-specific. Spike-specific CD8 T cells were identified based on activation-dependent co-expression of CD137 and IFNγ and CD107a and IFNγ, respectively. At least a twofold increase in frequency after stimulation compared with unstimulated controls was defined as responder.

### Analytical methods

All flow cytometric analyses were performed using a BD FACS Fortessa (BD Biosciences, Franklin Lakes, NJ, USA). To ensure comparable mean fluorescence intensities (MFIs) over time of the anaylses, Cytometer Setup and Tracking beads (CST beads, BD Biosciences, Franklin Lakes, NJ, USA) and Rainbow Calibration Particles (BD Biosciences, Franklin Lakes, NJ, USA) were used. For flow cytometric analysis, the following fluorochrome-labeled antibodies were used: BUV737 anti-CD11c (BD, clone B-ly6), BUV395 anti-CD14 (BD, clone M5E2), BUV395 anti-CD3 (BD, clone UCHT1), BV786 anti-CD27 (BD, clone L128), BV711 anti-CD19 (BD, clone SJ25C1), BV605 anti-CD24 (BD, clone ML5), BV510 anti-CD10 (BD, clone HI10A), BV421 anti-CXCR5 (BD, clone RF8B2), PE-Cy7 anti-CD95 (ThermoFischer, Waltham, MA, USA clone APO-1/Fas), PE-CF594 anti-IgD (Biolegend, San Diego, CA, USA, clone IA6-2), APC-Cy7 anti-CD38 (Biolegend, clone HIT2), PE-Cy7 anti-IgG (BD, clone G18-145), anti-IgA-Biotin (BD, clone G20-359), BV650 anti-IgM (BD, clone MHM-88), FITC anti-HLA-DR (Biolegend, clone L234), PE anti-CD21 (BD, clone B-ly4), APC anti-CD22 (BD, clone S-HCL-1), FITC anti-TNFα (Biolegend, clone Mab11), BV650 anti-IFNγ (BD, clone 4S.B3), BV786 anti-CD40L (Biolegend, clone 24-31), PE-CF594 anti-CD137 (Biolegend, clone 4B4-1). Number of absolute B cells was measured with Trucount (BD) and samples were processed according to the manufacturer's instruction.

**Sorting of plasmablasts, B and T cells from peripheral blood for single cell analysis**

As reported [3], cells were enriched from peripheral blood using StraightFrom® Whole Blood CD19, CD3 and CD138 MicroBeads (Miltenyi Biotec) according to manufacturer's instructions, incubated with Fc Blocking Reagent (Milteniy Biotec) and subsequently stained up to $5x10^6$ cells per 100µL with the following anti-human antibodies: VioBlue anti-CD3 (Miltenyi Biotec, clone BW264/56), VioBlue ant-CD14 (Miltenyi Biotec, Clone TÜK4), VioBlue anti-CD16 (Miltenyi Biotec, clone REA423), PE anti-CD27 (Miltenyi Biotec, clone MT271), APC anti-CD38 (BioLegend, clone HIT2). Sorted populations were identified as plasmablasts (DAPI-CD3-CD14-CD16-CD38++CD27++), memory B cells (DAPI-CD3-CD14-CD16-CD38-CD27+) and activated T cells (DAPI-CD3+CD14-CD16-CD38++HLA-DR+). The three sorted populations were pooled and further processed for single cell RNA sequencing.

**Single Cell RNA-library preparation and single-cell transcriptome sequencing**

Sequencing was performed on a NextSeq500 device (Illumina) using High Output v2 Kits (150 cycles) with the recommended sequencing conditions for 5' GEX libraries and as reported [3]. In particular, transcriptome profiles were merged, normalized, variable genes were detected and a Uniform Manifold Approximation and Projection (UMAP) was performed in default parameter settings using FindVariableGenes, RunPCA and RunUMAP with 30 principle components. Expression values are represented as ln (10,000 * UMIsGene) /UMIsTotal +1). Transcriptionally similar clusters were identified using shared nearest neighbor (SNN) modularity optimization, SNN resolutions ranging from 0.1 to 1.0 in 0.1 increments were computed, or gating was performed manually using the Loupe Browser (10x Genomics). Data from transcriptome and immune profiling were merged by the same cellular barcodes.

**Data Analysis and Statistics**

All samples included in the final analyses had at least 1 × $10^6$ events with a minimum threshold for CD19+ cells of 5,000 events apart from RTX patients: minimal recorded CD19+ events in the RTX group were 16 and 45 events respectively, out of > 1Mio total recorded events. Flow cytometric data were analyzed by FlowJo software 10.7.1 (TreeStar, Ashland, OR, USA). GraphPad Prism Version 5 (GraphPad software, San Diego, CA, USA) was used for statistical analysis. For multiple group comparison, two-way ANOVA with Šidák's post-test for multiple comparison or Kruskal-Wallis with Dunn´s post-test was used. Spearman correlation coefficient was calculated to detect possible associations between parameters or disease activity, respectively. P-values < 0.05 were considered significant. Data on cellular subsets of one RTX patient with concomitant chronic lymphocytic leukemia, was excluded in the figures 2, 3 and 4. For one RTX patient and 6 HC there was not enough material to perform FACS T cell staining for activated T cells (Fig. 3 B and C). Correlation matrix was calculated using base R and corrplot package (R Foundation for Statistical Computing) using the Spearman method (n=17, one excluded with concomitant CLL, one because of partially missing T cell data).

**Supplementary figures.**

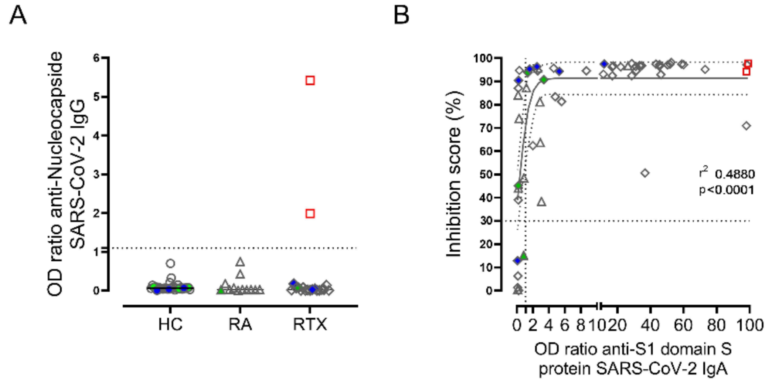


**Figure S1: Nucleocapside among all donors.**

(A) Humoral immune response against SARS-CoV-2 was assessed by ELISA for nucleocapside in HC (n=30), RA (n=12) and RTX (n=19) 6±3 days after 2nd SARS-CoV-2 vaccination. (B) Correlation between IgA titers and neutralizing antibodies. Kruskal-Wallis with Dunn´s post-test. *p<0.05, **p<0.01, ***p<0.001, ****p<0.0001. Color code: previously infected individuals are indicated as red quadrats; 2x vaccinated with ChAdOx1 indicated in green; 2x heterologous vaccinated 1x ChAdOx1 followed by 1x BNT162b2, indicated in blue.

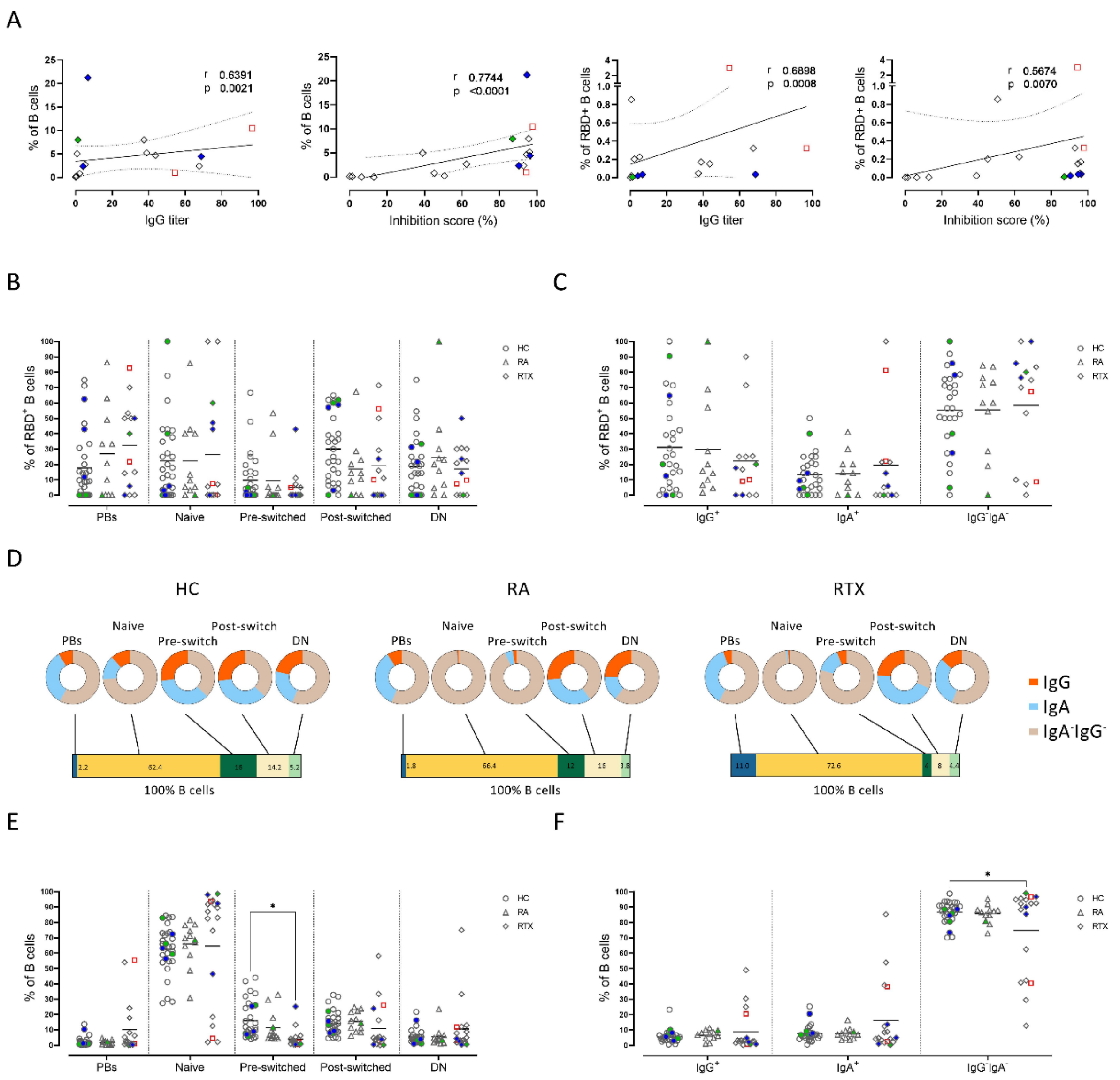


**Figure S2: Subset distribution of B cells and RBD+ B cells of HC, RA and RTX patients**

(A) Correlation between B cell frequencies and RBD+ frequencies with IgG titers, as well as neutralizing antibodies. (B) Frequencies of RBD+ B cells naïve, pre-switch memory, post-switched and double negative B cells (as described in Figure 2A) in HC (n=28), RA (n=11) and RTX (n=14). Only donors with detectable RBD+ B cells were included. (C) Frequencies of IgA+, IgG+ and IgA-IgG- RBD+ B cells 6±3 days after 2nd vaccination in HC, RA, and RTX. (D) Frequencies of plasmablasts, naïve, pre-switch, post-switched and double negative B cells (bar) and immunoglobulin isotype distribution among subsets (cakes) in HC, RA and RTX patients, 6±3 days after $2^{nd}$ vaccination in HC (n=30), RA (n=12) and RTX (n=18). (E) Frequencies of RBD+ B cells naïve, pre-switch memory, post-switched and double negative B cells (as described in Figure 2A) in HC, RA, and RTX. (F) Frequencies of IgA+, IgG+ and IgA- IgG- RBD+ B cells 6±3 days after 2nd vaccination in HC, RA, and RTX. *p<0.05, **p<0.01, ***p<0.001, ****p<0.0001. Color code: previously infected individuals are indicated as red quadrats; 2x vaccinated with ChAdOx1 indicated in green; 2x heterologous vaccinated 1x ChAdOx1 followed by 1x BNT162b2, indicated in blue.

Fig S3

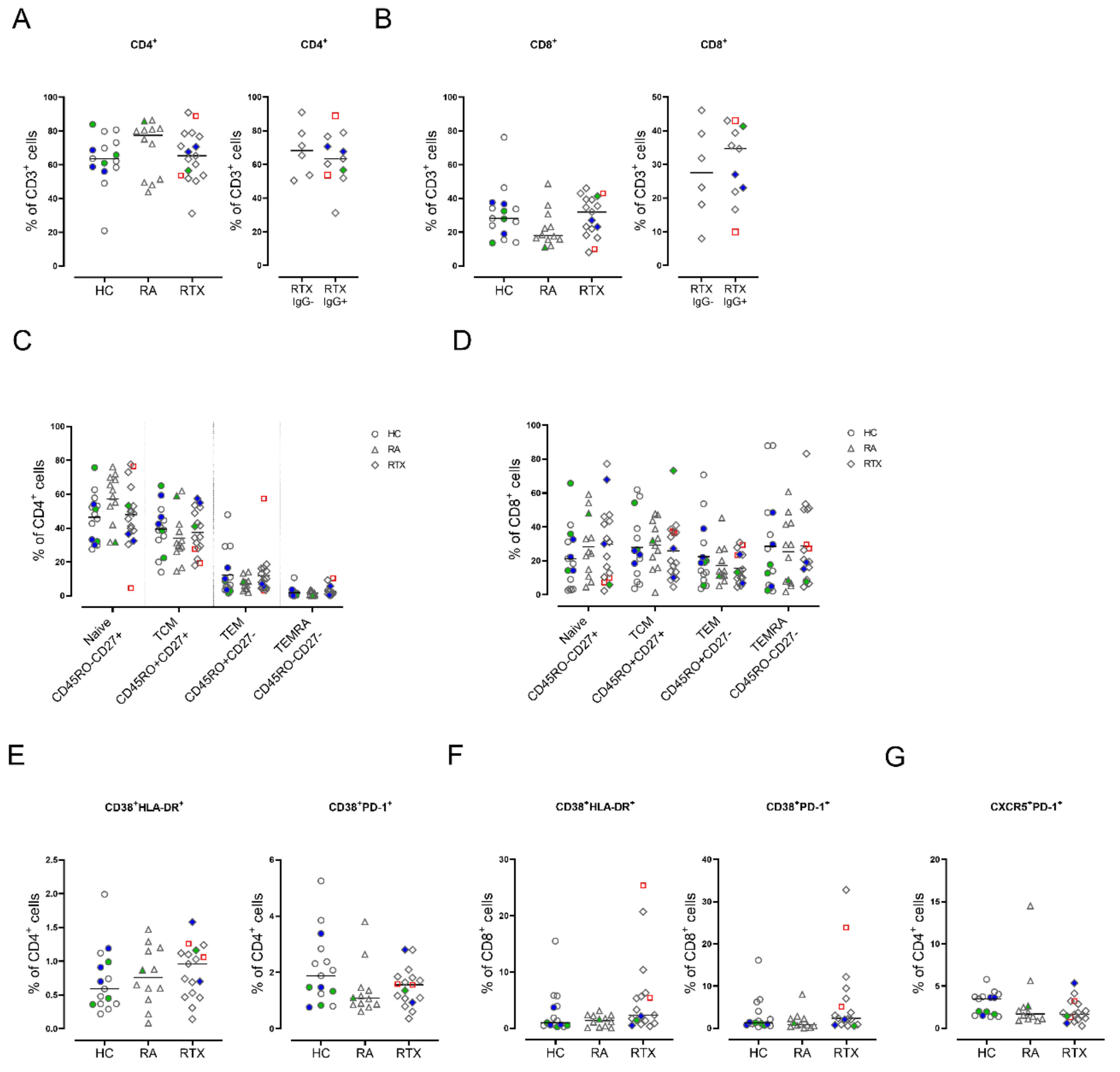


**Figure S3: Subset distribution of T cells in HC, RA and RTX.**

(A-B) Frequencies of (A) CD4 and (B) CD8 T cells in HC (n=14), RA (n=12) and RTX (n=17), and in RTX IgG+ (n=11) compared to RTX IgG- (n=6) 6±3 days after $2^{nd}$ vaccination. (C-D) Subset distribution of (C) CD4 and (D) CD8 T cells according to CD45RO/CD27 expression among HC, RA, and RTX. (E-F) Frequencies of activated CD38+HLA-DR+ and CD38+PD-1+ in (E) CD4 and (F) CD8 T cells in HC, RA, and RTX. (G) Frequency of circulating TfH-like CD4 T cells in HC, RA and RTX. (A-B) Mann Withney test. (A-B, E-G) Kruskall Wallis with Dunn´s post-test. (C-D) Two way ANOVA with Šidák post-test. Interaction effect was not significant. *p<0.05, **p<0.01, ***p<0.001, ****p<0.0001.

Color code: previously infected individuals are indicated as red quadrats; 2x vaccinated with ChAdOx1 indicated in green; 2x heterologous vaccinated 1x ChAdOx1 followed by 1x BNT162b2, indicated in blue.

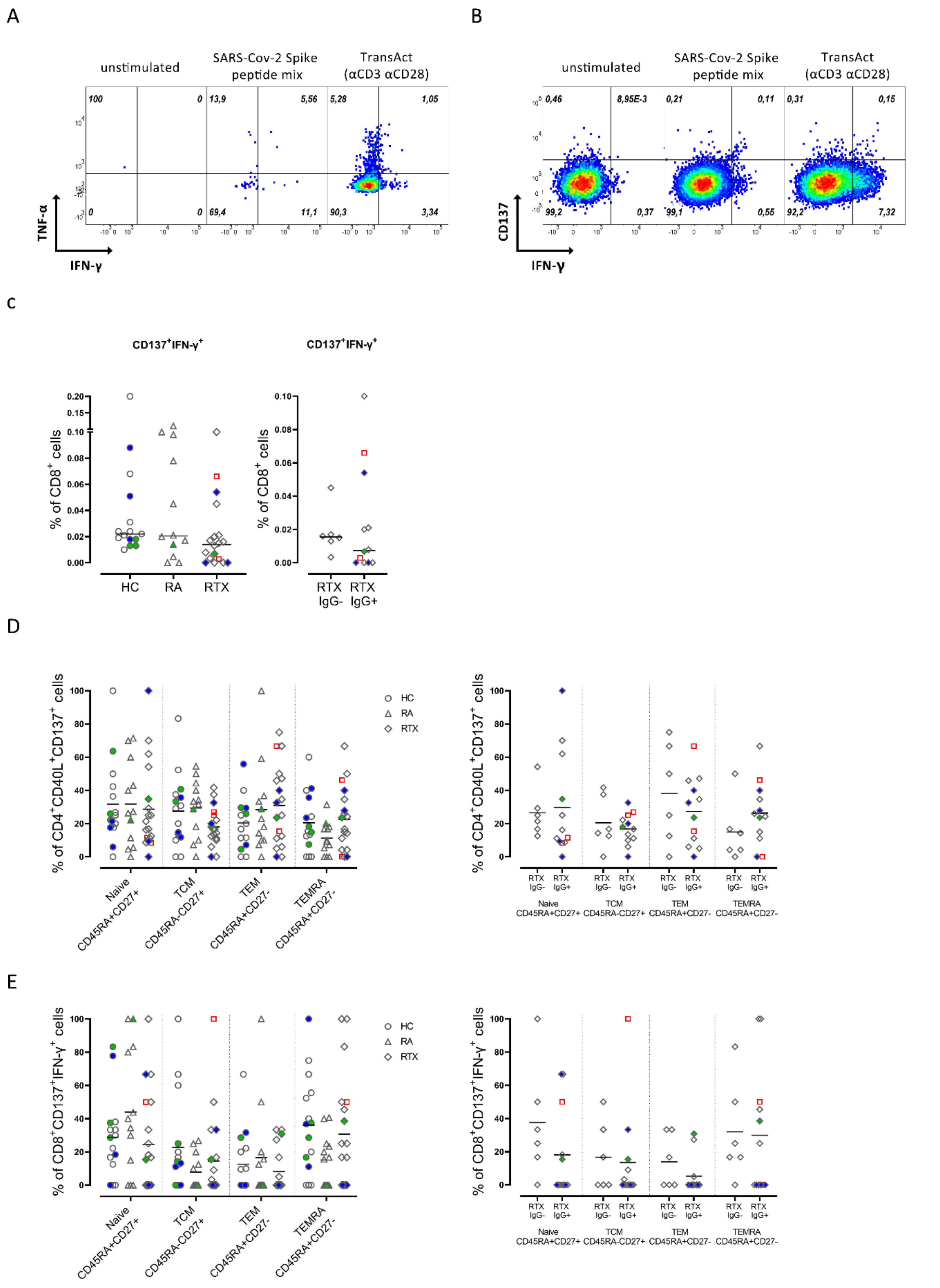

**Figure S4: Subset distribution of antigen-specific CD4 and CD8 T cells in HC, RA and RTX.**

(A) Representative plots of TNFα and IFNγ expression of spike-specific CD4 T cells after culture of PBMC unstimulated (left) or stimulated with SARS-CoV-2 Spike peptide mix (center) and TransAct (right) as described in materials. (B) Representative plots for antigen-specific CD8 T cells (CD137+IFNγ+) after culture of PBMC unstimulated (left) or stimulated with SARS-CoV-2 Spike peptide mix (center) and TransAct (right) . (C) Frequency of antigen-specific CD8+CD107a+IFNγ+ T cells in HC (n=15), RA (n=12, and RTX (n=18). (D-E) Subset distribution of (D) spike-specific CD4 T cells and (E) CD8+CD137+IFNγ+ T cells after peptide stimulation according to CD45RA/CD27 expression in HC, RA, and RTX, and in RTX IgG+ (n=11) compared to RTX IgG- (n=6) 6±3 days after 2$^{nd}$ vaccination. (D-E) Two way ANOVA with Šidák post-test. Interaction effect was not significant. *p<0.05, **p<0.01, ***p<0.001, ****p<0.0001. Color code: previously infected individuals are indicated as red quadrats; 2x vaccinated with ChAdOx1 indicated in green; 2x heterologous vaccinated 1x ChAdOx1 followed by 1x BNT162b2, indicated in blue.

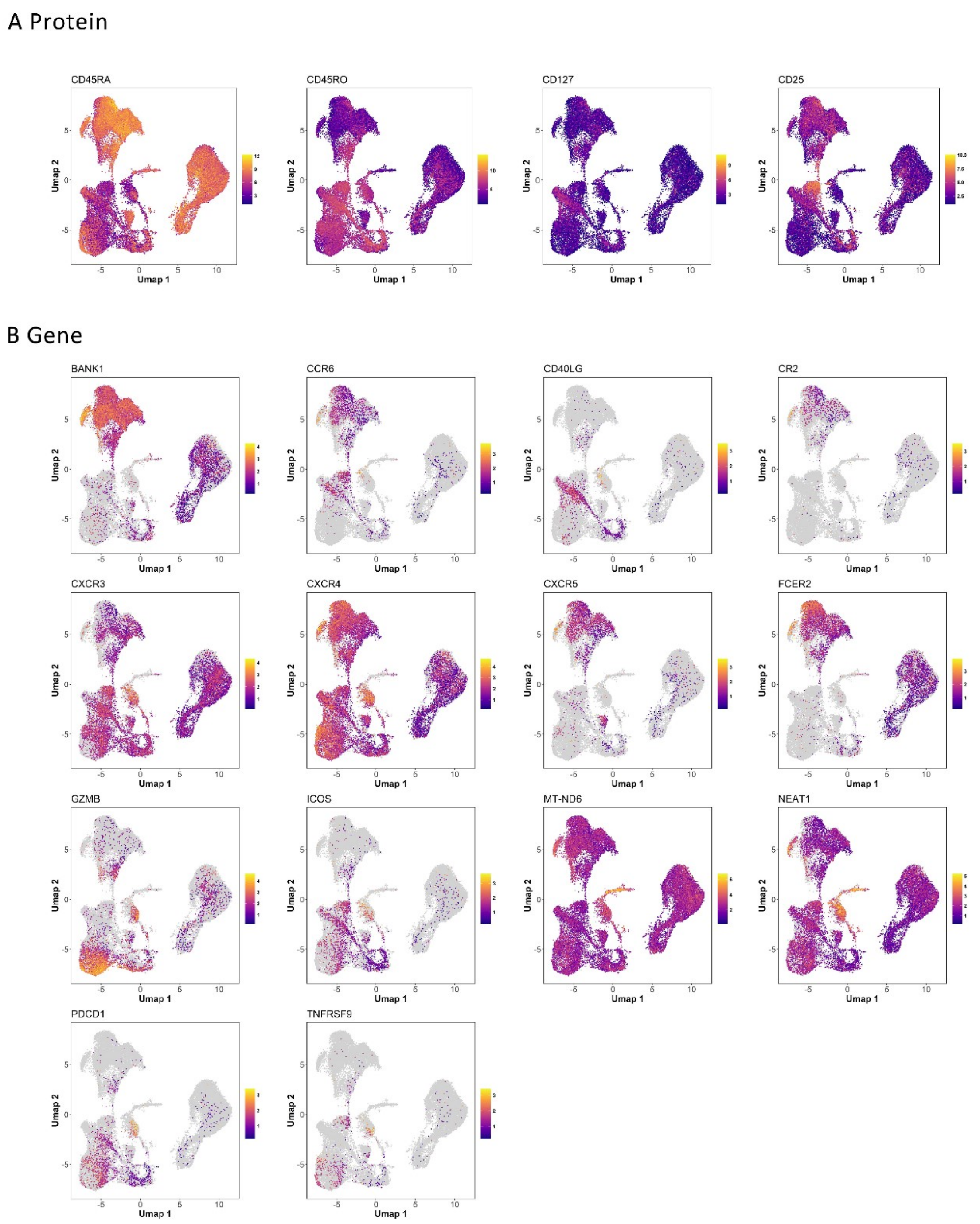


**Figure S5: Selected signature genes and protein level visualized after single cell transcriptome and CITE Seq analyses**

UMAP representation of the expression levels of selected signature protein level (A) and genes (B) for pooled sequenced cells. UMAP clustering as described for Fig. 5C.

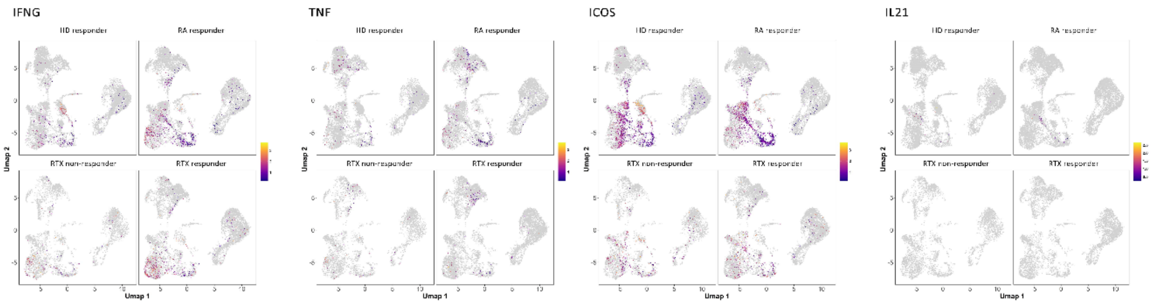


**Figure S6: UMAP representation of the expression levels of IFNG, TNF, ICOS, and IL21 for pooled sequenced cells.**

UMAP representation of the expression levels of *IFNG*, *TNF*, *ICOS,* and *IL-21* in HC, RA, IgG non-responder and responder RTX. UMAP clustering as described for Fig. 5C.

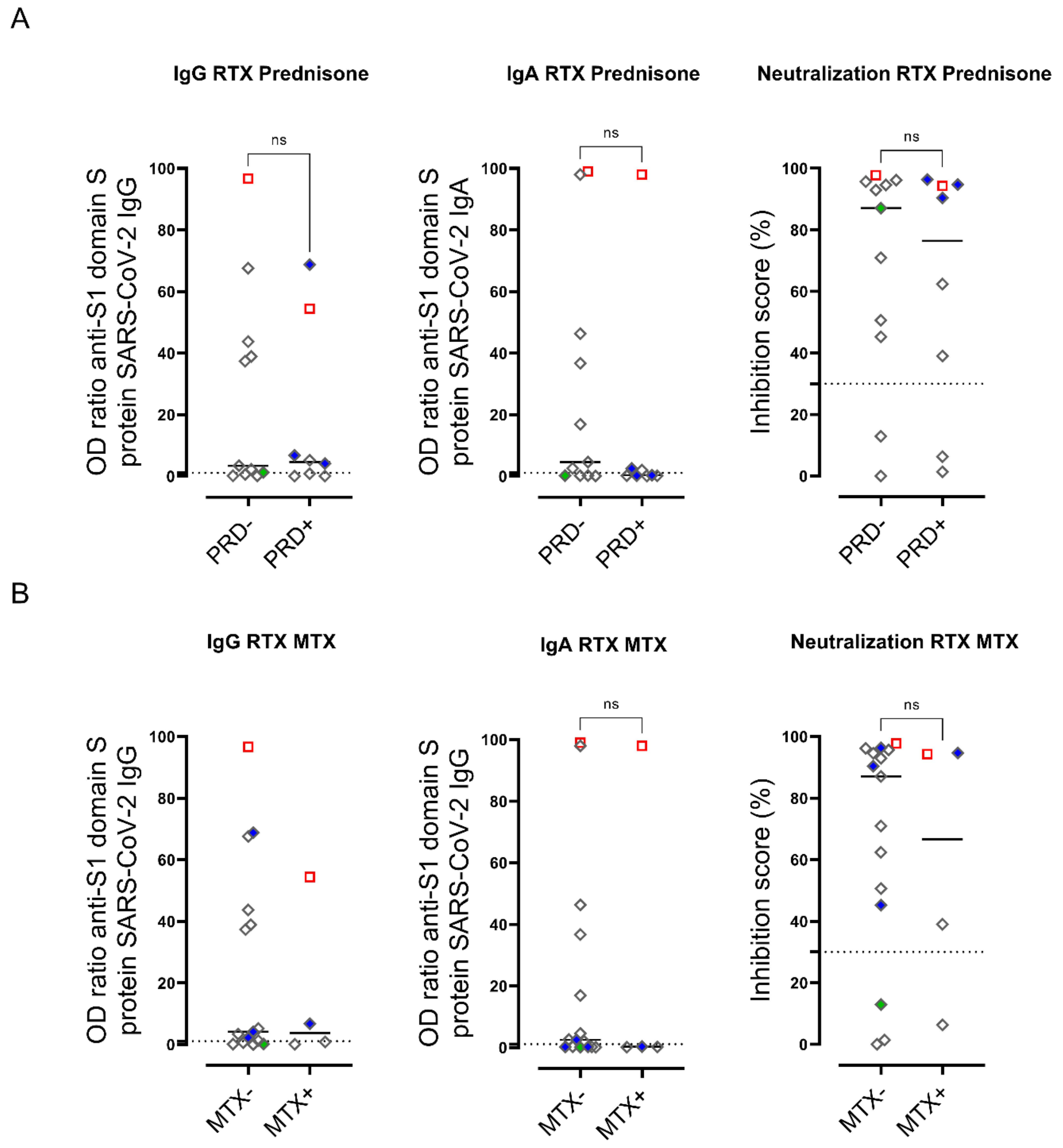


**Figure S7: Co-medication with Prednisolone or Methotrexate do not affect humoral response in RTX treated patients.**

Humoral immune response against SARS-CoV-2 was assessed by ELISA for spike protein S1 IgG, spike protein S1 IgA and virus neutralization by a blocking ELISA in RTX (n=19) 6±3 days after 2nd SARS-CoV-2 vaccination and treated or not with (A) Prednisolone (PDN, n=8) and Methotrexate (MTX, n=4). (A-B) Mann Withney test. *p<0.05, **p<0.01, ***p<0.001, ****p<0.0001. Color code: previously infected individuals are indicated as red quadrats; 2x vaccinated with ChAdOx1 indicated in green; 2x heterologous vaccinated 1x ChAdOx1 followed by 1x BNT162b2, indicated in blue.
